# Establishment of ‘natural antibodies’ during primary dengue infection

**DOI:** 10.1101/2025.08.09.25333157

**Authors:** T.W. Verdonckt, A.S. Vermeersch, C. Struyfs, M. Sahulčík, F. Van Nieuwerburgh, A. Waickman, O. Lagatie, K. K. Ariën

## Abstract

Dengue virus (DENV) poses a major global health burden, with limited vaccine availability and concerns that immunization of dengue-naïve individuals may exacerbate disease severity due to antibody-dependent enhancement. This challenge highlights the need for deeper insight into primary immune responses. Using a controlled human challenge model, we conducted a longitudinal analysis of B-cell receptor repertoire dynamics during primary DENV1 infection. Our study reveals that the acute-phase response is dominated by memory-derived B-cell clones, indicating pre-existing cross-reactive immunity. Concurrently, we identified highly public, convergent B-cell clones arising from naïve B cells, characterized by shared sequence features across individuals. These clones exhibit atypical maturation kinetics: while they switch to the IgG isotype during the convalescent phase, they retain germline-like sequences with limited somatic hypermutation. This suggests that affinity maturation is delayed compared to canonical responses. Our findings provide mechanistic context for previous reports of natural antibody responses in dengue and refine current models of how neutralizing and potentially enhancing antibodies emerge following primary infection. By resolving the timing and origin of early B-cell responses, this work contributes to a more precise framework for understanding flavivirus immunity. These insights may guide vaccine design strategies that aim to elicit protective immunity without enhancing the risk of severe disease.

**One Sentence Summary:** Primary DENV1 infection elicits convergent clones that undergo atypical maturation.

## INTRODUCTION

Dengue fever, a mosquito-borne disease caused by the dengue virus (DENV), remains one of the most pressing global health concerns, particularly in tropical and subtropical regions where the disease burden is most severe. With an estimated 400 million infections annually, dengue continues to affect vast populations, often leading to significant morbidity and mortality (*1*).

The virus exists as four distinct serotypes, DENV1 through DENV4 (*2*), each capable of causing a spectrum of illness. While most infections are asymptomatic (*3*), the disease can progress from mild febrile to severe manifestations such as dengue haemorrhagic fever (DHF) and dengue shock syndrome (DSS). Generally, disease progression follows three stages:

1. Febrile phase: characterized by high fever, rash, myalgia, and elevated viremia.
2. Critical phase: occurs in a subset of patients, typically around the time of defervescence and when antibodies become detectable, indicating immune control. This stage is marked by increased vascular permeability, thrombocytopenia, and risk of shock.
3. Recovery phase: clinical symptoms resolve as vascular permeability normalizes. This phase is associated with immune system stabilization rather than direct antiviral activity (*4–6*).

Severe dengue is defined by the development of one or more of the following features during the critical phase: severe plasma leakage leading to shock; fluid accumulation with respiratory distress; severe bleeding; and/or severe organ impairment (*6*). Despite extensive research, the underlying mechanisms of progression to severe dengue remain complex and elusive.

Importantly, the clinical outcome of DENV infections can be influenced by the host’s immune status. It has long been accepted that primary infections usually result in mild disease and that secondary heterologous infections carry a higher risk of severe outcomes, primarily due to immune mechanisms such as antibody-dependent enhancement (ADE) and original antigenic sin (OAS) (*7*, *8*). However, this paradigm is now being reevaluated. Recent evidence from pediatric cohorts in India (*9*) demonstrates that primary dengue infections can also lead to severe disease at comparable frequencies to secondary infections. These findings underscore that both primary and secondary infections can contribute significantly to dengue pathology. They also highlight the central role of the immune response, particularly B-cell-mediated mechanisms, in modulating disease severity.

Extensive efforts have aimed to clarify how immunity differs between primary and secondary DENV infections, especially given the known implications of ADE and OAS for severe disease.

During primary DENV infections, B-cells are activated and differentiate into antibody producing plasmablasts. Initially, IgM antibodies dominate, followed by a switch to IgG and IgA, both of which can contribute to viral clearance (*10*, *11*). This antibody response tends to be serotype-specific, effectively neutralizing the infecting DENV serotype but offering only limited cross-protection against heterologous serotypes. Germinal centre reactions involving T follicular helper (Tfh) cells further improve neutralizing capacity by driving antibody maturation and affinity increases (*12*).

In contrast, secondary infections rapidly reactivate memory B-cells, resulting in a massive plasmablast expansion that can account for up to 30% of circulating lymphocytes. These plasmablasts produce large amounts of IgG antibodies, which frequently exhibit cross-reactivity to multiple DENV serotypes but vary in neutralizing potential (*13*). However, memory B-cells originating from a prior DENV infection can preferentially produce antibodies against the original infecting serotype, a phenomenon known as original antigenic sin (*14*, *15*). Sub-neutralizing antibodies from such cross-reactive responses can facilitate viral entry into Fc receptor-bearing cells (e.g., monocytes, dendritic cells), thereby enhancing viral replication (*16*).

Dengue infection induces the emergence of highly public, convergent complementary determining region 3 (CDR3) heavy-chain amino acid motifs, such as ARQ(F/W/L/I/S)GNWFD(S/L/P) and AR(L/I)DYYYYYGMD(I/V/L) that are frequently observed during acute infection, particularly in secondary dengue cases (*10*, *17–19*). Parameswaran et al. (*18*) initially proposed that these convergent sequences arise from affinity-matured memory B-cells given their high mutation rates. Subsequent studies revealed that some convergent clones^1^ exhibit low somatic hypermutation (SHM) levels, suggesting an origin in naïve B-cells (*19*, *20*).

Notably, infection can also broadly affect the BCR SHM rates in antibody secreting cells (ASCs): primary dengue patients show reduced overall IgG SHM compared to healthy individuals, particularly during the acute phase of primary infections (*10*, *19*).

In light of these observations, Godoy-Lozano et al. (*19*) proposed a model where the initial IgG mediated response to dengue involves an innate-like antiviral mechanism that enables T-cell independent, extrafollicular differentiation of naive B-cells into ‘natural antibody’ secreting cells. These ASCs bear near-germline V(D)J genes; target conserved viral structures like capsids with relatively low affinity; and are often highly cross-reactive (*21*, *22*). They have been implicated in the early response to many human viruses including dengue (*19*, *23*). Because of their cross-reactive nature and minimal SHM, such antibodies might play a key role in ADE and OAS during secondary dengue. They may also act as valuable prognostic biomarkers for disease progression but present challenges for vaccine development.

Despite these advances, there remains an urgent need to better understand how ‘natural antibodies’ originate during primary DENV infections and how they influence disease outcome. Addressing these gaps is particularly relevant given the risks of ADE and OAS, both of which hinge on nuanced antibody dynamics.

In this study, we leverage longitudinal samples from a controlled human challenge model with an under-attenuated DENV1 strain (45AZ5) (*24*). By performing bulk B-cell receptor (BCR) sequencing and detailed immunoprofiling, we aim to characterize the ontogeny and potential clinical significance of natural antibodies elicited in primary dengue infections. The controlled nature of this model and the longitudinal experimental design offer unprecedented insights into the early B-cell response and its subsequent evolution.

## RESULTS

### Primary DENV1 infection induces individual- and isotype-specific immune activation

BCR sequencing libraries were generated from 1 µg RNA of each sample. As the lymphocyte count in peripheral blood can vary considerably between subjects and clinical conditions (*25*, *26*), we expected the unique sequence count of the samples to likewise differ considerably between study participants and timepoints. The total count of unique BCR heavy-chain sequences is shown in Supplementary Table 1. Samples day 0 of participant 6; and day 28 of participant 1; were not processed as extracted RNA quantity did not meet the library preparation input criteria.

Earlier studies in DF/DHF patients reported that DENV infections can trigger the massive release of virus-specific IgG1 secreting plasmablasts into the peripheral blood at days 4-7 after fever onset, especially in secondary infections (*11*, *13*, *17*). In these studies, the abundance of virus-specific IgA secreting cells was reported as 100 times lower. Interestingly, primary infections also induced an increased abundance of virus-specific IgM secreting cells (*13*). This timeframe approximates to days 13-16 of this study, with some variance between the study participants. Indeed, as reported previously, day 14 was characterized by a strong lymphocyte-activation associated gene module, increased IGH count, and high DENV-specific IgM titers, soon followed by DENV-specific IgA on day 16 and DENV-specific IgG around day 20 (*24*).

To verify if a similar increase of ASCs could be detected in our data, the count of unique IGH sequences of each isotype at each timepoint was plotted, normalized towards the mean values of days 0, 8, and 10 (Figure 1A). The plot shows that the increase of IGH sequence counts is highly subject dependent. Participants 4, 5, and 7 display a strong increase (> 2.5x) of IGHG, IGHM, and IGHA sequences on day 14 compared to earlier timepoints; whereas for participant 9 there is a moderate increase (∼2x) in the IGHG and IGHA sequence counts but a decrease (0.23x) in IGHM counts, hinting at a secondary immune response. No substantial increase in IGHA or IGHG counts is observed for participants 1, 2, 3, 6 and 8. Furthermore, we did not observe clear trends in the clonal expansion/evenness scores of the repertoires, except for a significantly decreased diversity of IGHM on day 14 relative to day 0 in 7 out of 9 participants (Supplementary Figure 1A).

**Figure 1.**
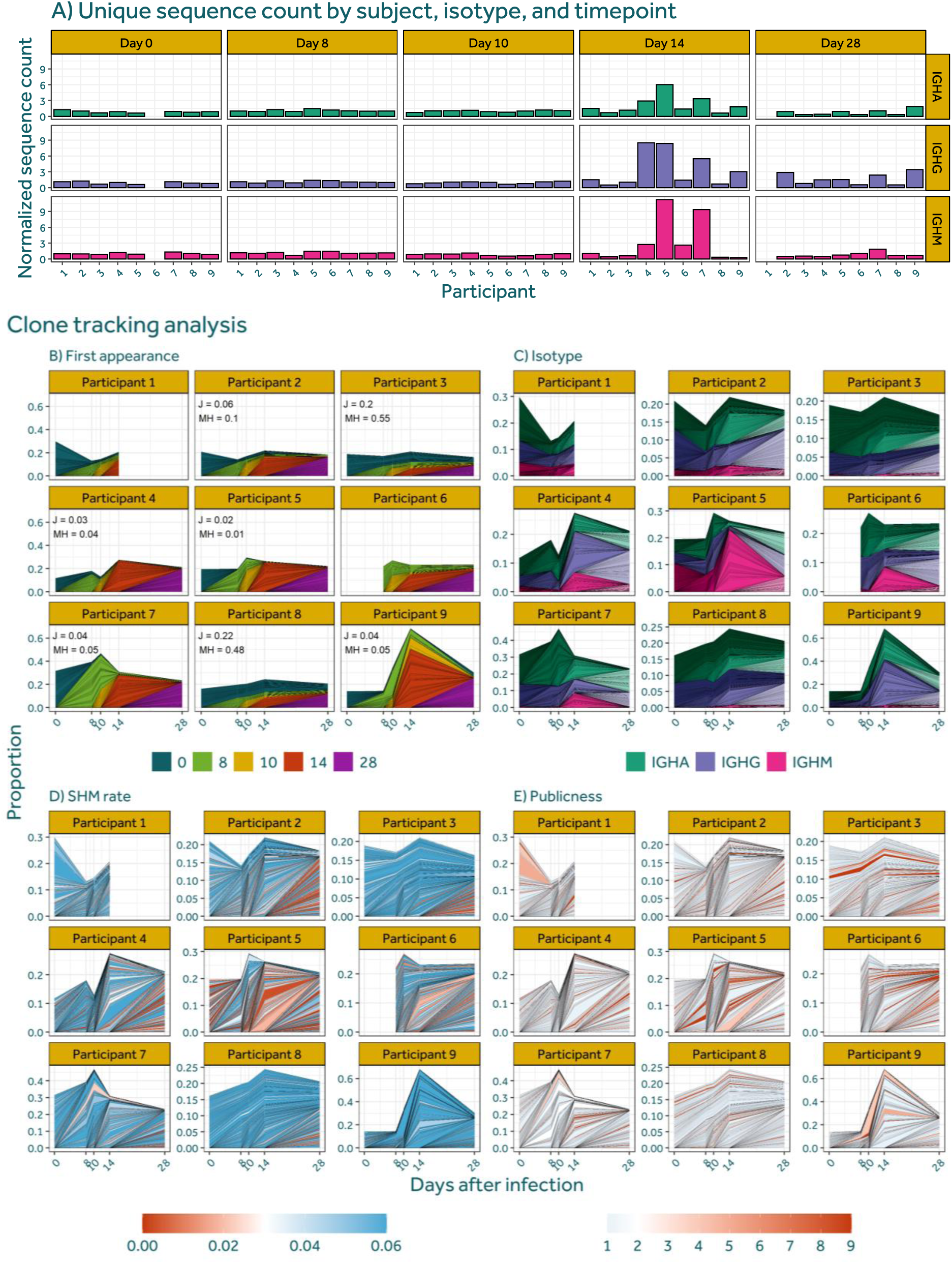
Sequence count and clone tracking analysis. (**A**) Count of unique IGH sequences normalized towards the mean of days 0, 8, and 10 for each participant. (**B**) Clone tracking plot with mean Jaccard (J) and Morisita-Horn (MH) values, the clones are coloured by timepoint of first appearance; (**D**) the rate of somatic hypermutation in the v-gene; (**E**) or their publicness. The clones were then split, grouped, and coloured by isotype (**C**) and ranked as well as shaded by timepoint of first appearance. In plot (B) the y-axis is shared for all participants to allow direct comparison of the degree of clonal expansion.

To capture the dynamics of the immune repertoires, a clone-tracking analysis was conducted. Clonal abundances were first estimated for each participant, after which the 200 most abundant clones at each timepoint were plotted and mean repertoire overlap metrics computed (Figure 1B). Participants 4, 5, 7, and 9 have a high clonal turnover rate, with Morisita-Horn (MH) indices of 0.05 or less, thereby indicating a robust adaptive immune response. Participant 2 displayed a weaker clonal turnover with a MH value of 0.1, whereas participants 3 and 8 had MH values of approximately 0.5 and retained top clones that were stable over time. Participants 1 and 6 cannot be classified due to missing timepoints. These findings align with previously published data on cellular proliferation and the differentially expressed gene module for lymphocyte activation of these samples (*24*). Analysis of isotype proportions over time (Figure 1C) reveals that in participants 2, 3, and 8, the ratio between isotypes remains stable throughout the study period. In contrast, participants 4, 5, 6, and 7 exhibit a marked increase in the IGHM fraction on day 14, suggesting the expansion of naïve clones and the initiation of an immune reaction in response to the DENV1 challenge. Notably, this IGHM expansion is most pronounced in participant 5, for whom the highest virion- and non-structural 1 (NS1)-binding IgM titres were observed (*24*).

Next, we examined the relationship between clonal expansion and somatic hypermutation. The SHM rate of the clones plotted in Figure 1D align closely with the partition of participants also observed through repertoire turnover and the lymphocyte activation differential gene expression (DGE) module (*24*). Specifically, participants 2, 3, and 8 display a sustained high SHM rate in their largest clones through day 14 (consistently above 0.025), followed by an emergence of less mutated clones at timepoint 28. This temporal pattern suggests that B-cells with considerable levels of affinity maturation are dominant during the acute phase of infection, while naïve-derived lower-SHM clones only appear in the convalescent phase. In contrast, participants 4, 5, 6, and 7 show no apparent time-dependent trend in SHM, as lower-SHM clones are present to the same extent at all sampled intervals. Finally, participant 9 exhibits a distinctive pattern wherein its highly expanded clones maintain a consistently elevated SHM rate across every timepoint, irrespective of when the clones emerge or the degree of their expansion, suggesting that the immune response of participant 9 was dominated by memory-derived B-cells. The repertoire of participant 1 could not be classified due to missing the sample from timepoint 28

To assess the extent of convergent evolution in the largest clones of each participant, the publicness of the expanded clones was assessed. Public clones were defined as those clones that contain at least one sequence belonging to a ‘public cluster’. The latter are generated by pooling the repertoires of all participants and clustering the dataset into sets of sequences with the same V-J gene pairing and 0.2 length-normalized hamming distance threshold of the amino-acid junction sequence.

The ‘publicness’ of a clone is then determined by the highest number of participants from which sequences clustered with any of the sequences in the clone. Figure 1E illustrates that the degree of publicness among the most expanded clones remains largely unchanged by DENV1 infection. In other words, highly expanded clones do not exhibit an increased tendency to be shared across multiple participants when compared with pre-infection samples.

### Infection with DENV1 triggers the expansion of naïve, highly public clones with low mutation rate

Next, we aimed to better characterize the DENV1-specific immune response during primary infections. Previous studies have demonstrated that individuals exposed to the same antigen can develop public and antigen-specific CDR3 sequences (*27*). To investigate this phenomenon, we identified public IGH junction sequences, stratified by timepoint and isotype (Figure 2A).

**Figure 2.**
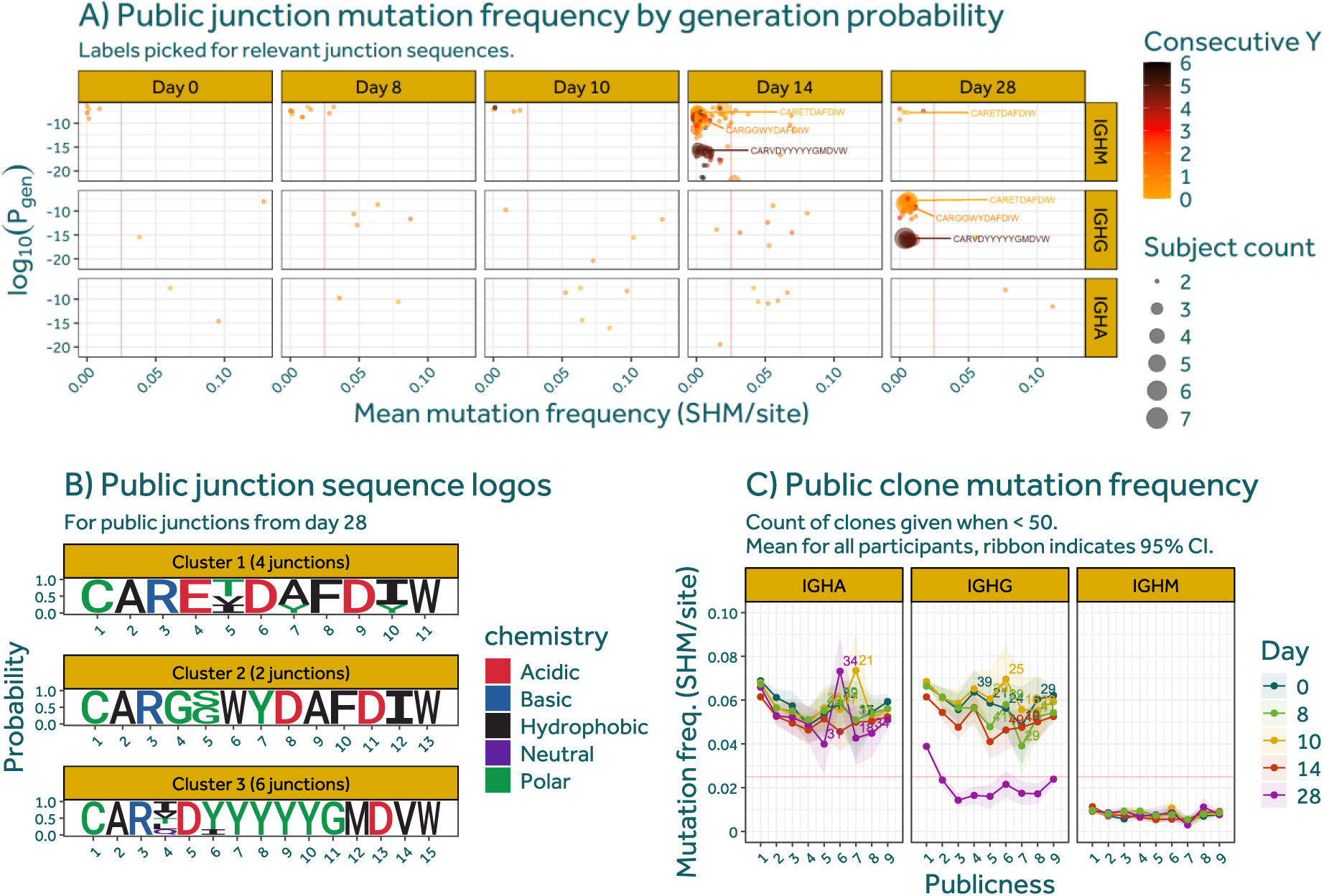
Emergence of public and naïve BCRs during primary DENV1 infection. (**A**) Analysis of public junctions in response to primary DENV1 infection. For each timepoint and isotype, junction amino acid sequences shared by two or more individuals are plotted. The size of the points indicates the number of subjects in which the junction is present. The color indicates the count of consecutive tyrosine residues. The mean SHM rate of all sequences with the public junction is plotted on the x-axis, and the junction generation probability as calculated through OLGA on the y-axis. Selected junction sequences are labelled. Vertical red line indicates 0.025 SHM rate threshold. (**B**) Sequence logos for clusters of public junction sequences 28 days after primary infection with DENV1. Junction sequences shared by three or more participants were clustered using a Levenshtein distance of 2. (**C**) Analysis of the correlation between the average SHM rate and the publicness of a clone, split by isotype and timepoint.

Public junction sequences can arise due to (i) a high intrinsic generation probability, (ii) the convergent selection of rare naïve junctions, or (iii) affinity maturation-driven convergent evolution (*28*). To distinguish between these mechanisms, we plotted generation probability (*p-gen*) on the y-axis and mean somatic hypermutation rate on the x-axis.

On days 0, 8, and 10, a baseline pattern can be observed where all public junctions are shared between only two participants. Public IGHM junctions are defined by a low SHM rate (generally under 0.025 mutations per site) and a relatively high generation probability (larger than 10^-10^), suggesting that these junctions are public due to their high generation probability from the germline. Public IGHG and IGHA junctions are characterized by a high SHM rate and lower generation probability, suggesting that these junctions are public through convergent evolution.

During the critical phase on day 14, a strong effect is observed for public IGHM junctions: the total count of public junctions increased significantly, with a large fraction of these junctions being shared by three or more individuals. The public junctions display a wide range of generation probabilities while maintaining a low SHM rate. This suggests that these junctions became public through convergent selection of the naïve BCR repertoire.

During the convalescent phase on day 28, the previously identified public junctions have undergone isotype switching towards IGHG. As predicted by Godoy-Lozano et al. (*19*), this occurred without an increase of the SHM rate, suggesting a T-cell independent maturation process.

Junctions of day 28 that are shared by three or more participants can be grouped into three clusters with a Levenshtein-distance of 2 (Figure 2B). Cluster 3 is characterized by a Y_5_ motif and has previously been described as a convergent signature of dengue infection, predominantly found in the acute stage of secondary infections (*18*). Tyrosine-rich motifs were also identified to play an important role in the immune response of mice immunized with dengue E-domain III (EDIII) and it is hypothesised that they enable the binding of antibodies to glycans of the E protein (*20*).

The probability of a junction being public is inversely proportional to the SHM rate (*29*). As such, the SHM values presented in Figure 2A might not be representative of the clones to which the public junction sequences belong, as the detected junctions might be biased towards less evolved branches of the phylogenetic tree.

To control for this bias and determine whether B-cells expressing public junctions undergo affinity maturation, we analysed the phylogenetic trees of sequences from the identified clusters (Figure 3).

**Figure 3.**
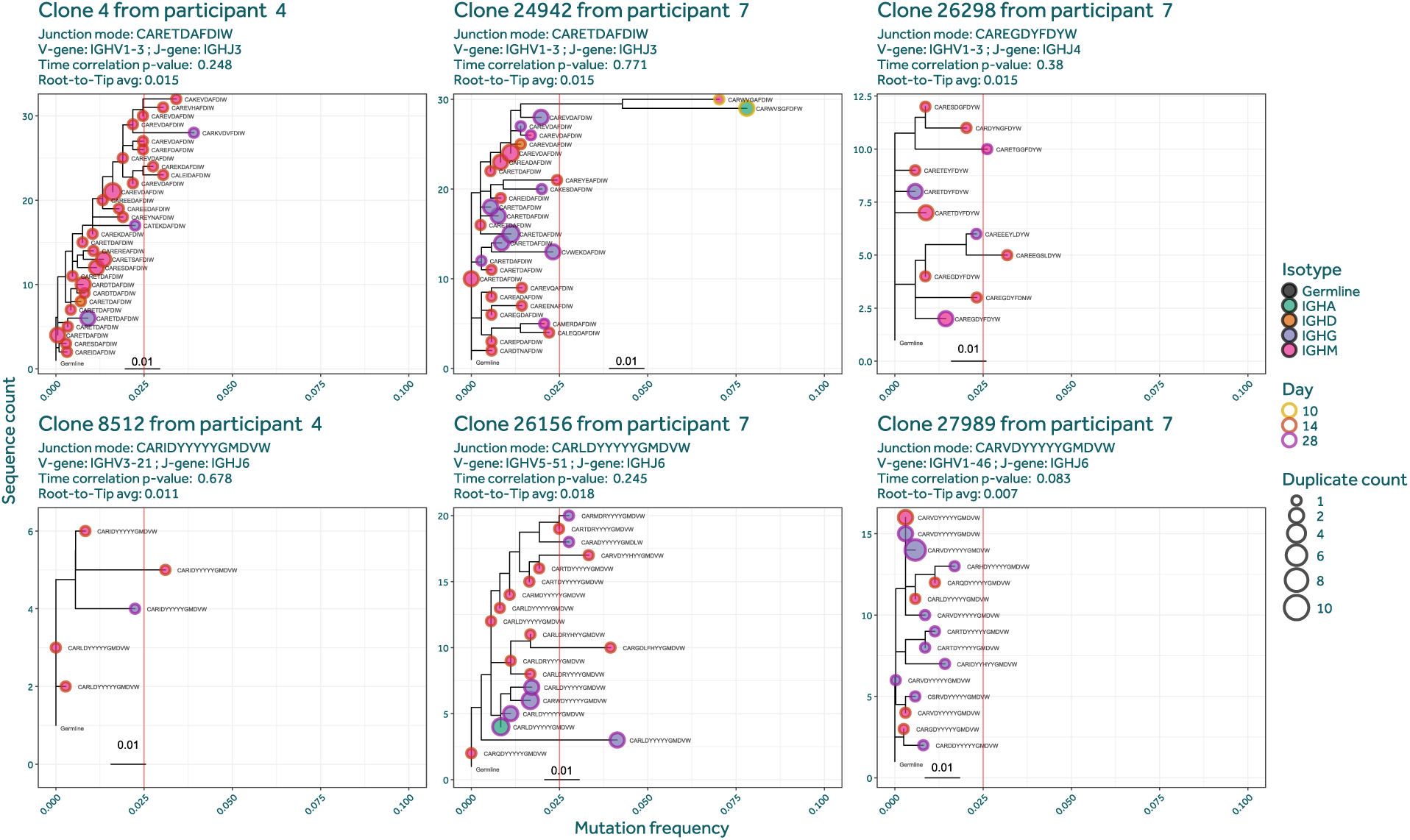
Phylogenetic trees of clones containing public junctions. Selection of phylogenetic trees from clones expressing public junction sequences belonging to cluster 1 (upper row) and 3 (lower row). Nodes coloured by isotype (fill) and timepoint (border). Node size represents duplicate count. Vertical red line indicates 0.025 SHM threshold.

The trees support the findings of Figure 2A that the public junctions first appeared on day 14 as IGHM before switching to predominantly IGHG on day 28. Importantly, the trees also demonstrate that the clones do not undergo affinity maturation within the timeframe studied, as the sequences almost exclusively maintain SHM rates under 0.025 up to day 28 after infection.

Identical public junction sequences are very rare, even between subjects exposed to the same antigen. In order to enhance analytical sensitivity, public clones were therefore defined. Figure 2C illustrates the association among a clone’s degree of ‘publicness’, its sampling timepoint, and its rate of somatic hypermutation.

This figure reveals an overall dynamic relationship between the average mutation frequency of IGHG/IGHA clones and their level of publicness. For publicness values 1 through 5, the mutation frequency is inversely proportional, whereas it stabilizes once higher publicness values are reached.

The figure further demonstrates that neither the SHM rate of IGHM sequences nor that of IGHA sequences changed with the sampling timepoint. In contrast, a pronounced effect was noted for IGHG sequences: up to 14 days post-infection, IGHG sequences showed SHM rates comparable to IGHA, which aligns with observations in healthy individuals (*30*). However, by day 28, the SHM rate in IGHG clones dropped sharply, remaining below 0.025 SHM/site for all public clones.

Natali et al. (*20*) reported increased CDR3 lengths in memory B-cells of patients infected with DENV2 and higher Shannon entropy per CDR3 length in mice immunized with DENV2 envelope than control mice or mice immunized with only EDIII. We replicated these analyses but found no temporal effect of primary DENV1 infection on either BCR property (Supplementary Figure 2).

## Primary DENV1 infection significantly alters the V-J gene-family pairing

We investigated the effect of clone expansion induced by DENV1 infection on the V–J gene-family pairing of immune repertoires. To this end, differences in V–J gene-family usage at days 14 and 28 post-infection were assessed relative to day 0 through a zero-inflated negative binomial mixed model with Benjamini–Hochberg correction. Changes were considered significant when they exceeded an absolute log2(fold-change) of 1 with a corrected p-value ≤ 0.05. Volcano plots were generated (Supplementary Figure 3), and data were visualized (Figure 4A).

**Figure 4.**
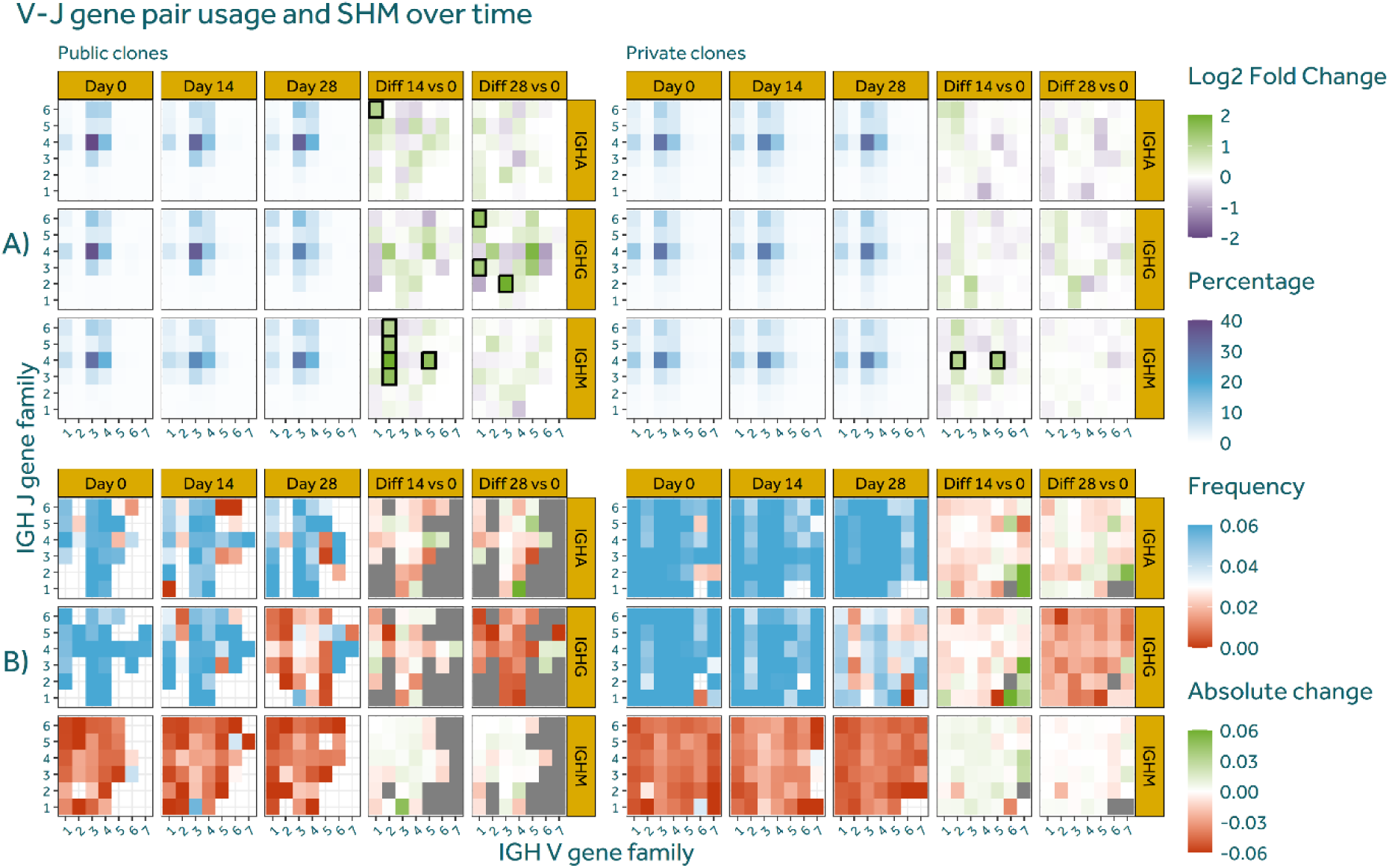
V-J gene pair usage and SHM over time. (**A**) Primary DENV1 infection significantly alters the relative abundance of V-J gene family pairing. Boxes indicate significant (zero-inflated negative binomial mixed-effects model with Benjamini–Hochberg correction p < 0.05 and |log2(fold change)| > 1) changes in gene-family pair abundance. (**B**) Primary DENV1 infection decreases the SHM rate of IGHG sequences.

Public clones displayed marked differences in gene-family usage between days 14 and 28. On day 14, most changes were detected in the IGHM isotype, where the V2 family paired more frequently with J3 through J6. Interestingly, only a single gene-family pair, V1–J6, showed a significant increase for IGHA. By day 28, alterations were limited to IGHG, characterized by upregulation of the V1–J6 pairing as well as V1–J3, and V3–J2. It was shown that the IGHJ6 gene family contributes substantially to the generation of tyrosine-rich CDRH3s in humans. The observed upregulation of V1–J6 for IGHG was also reported in the memory B-cell compartment of DENV2 patients (*20*).

In private clones, V2–J4 and V5–J4 were significantly elevated for IGHM on day 14 compared to day 0.

The results presented in Figure 2C and Figure 2E suggest that the DENV-specific immune response triggers a decrease in the SHM rate of IGHG sequences by day 28 after infection. To investigate whether this reduction is limited to specific V- or J- families, or a broader property of the repertoire, the SHM rate was plotted for each gene-family pair at timepoints 0, 14, and 28; split by isotype, timepoint, and between public/private clones.

In Figure 4B, IGHM sequences consistently exhibit low somatic hypermutation levels, irrespective of timepoint, gene pair, clone publicness, or differential expression between timepoints. By contrast, IGHA sequences maintain relatively high SHM rates overall, with moderate temporal fluctuations in specific gene pairs. A pronounced reduction in SHM is noted for IGHG at day 28, aligning with observations in Figure 2E. This decrease is more pronounced among public clones than private ones and inversely correlates with differential gene-pair expression (Supplementary Figure 4).

When differentially expressed gene family pairs were examined, the V1–J6 IGHA pairing on day 14 showed no change in SHM, whereas the corresponding IGHG pairing at day 28 displayed a marked reduction. This contrast supports the idea that IGHA and IGHG subsets emerge via distinct biogenetic pathways. Specifically, the higher SHM in IGHA indicates a memory-derived lineage, while the minimal SHM observed in IGHG is consistent with derivation from naïve IGHM clones, as suggested by Figure 2C.

## DISCUSSION

### Primary DENV1 infection induces participant- and isotype-specific immune activation

The DENV1-induced repertoire dynamics presented in this study can be classified into three groups based on the relative unique sequence counts and clone tracking results presented in Figure 1, and the lymphocyte activation scores previously reported by Waickman et al. (*24*).

The first group, composed of participants 2, 3, and 8, display no marked increase in unique sequence counts on day 14 for any isotype (Figure 1A), a low repertoire turnover (Figure 1B), and maintain isotype proportions in the dominant clones (Figure 1B). In this group, the SHM rate of expanded and novel clones predominantly decreased on day 28 relative to previous timepoints (Figure 1C). This suggests a moderate and delayed antibody-mediated immune response.

The second group, composed of participants 4, 5, 6, and 7, is marked by an increase of unique sequence counts for all isotypes on day 14, a strong repertoire turnover (mean MH ≤ 0.05), and a sharp expansion of IGHM dominant clones on day 14. In this group, the SHM rate of expanded clones does not show a temporal effect. The higher sequence count on day 14 is in agreement with an increase in peripheral blood plasmablasts and, given the increase of IGHA sequences, is especially relevant considering mounting evidence that IgA can play an important role in neutralizing DENV particles without inducing ADE and can even antagonize the ADE induced by IgG (*31–33*). Combined, these findings indicate a primary but stronger immune response compared to participants from group one that may be modulated by different B-cell maturation dynamics.

Lastly, participant 9 displays intermediate properties, marked by a strong increase in unique IGHA and IGHG sequences; a high repertoire turnover; and a rapid hyperexpansion of highly mutated clones at day 8 and 10 (Figure 1D supported by Supplementary Figure 1B and Supplementary Figure 5) that persist throughout the study period. However, this participant lacks a detectable IGHM response. These findings are indicative of a secondary, predominantly cross-reactive immune response, possibly caused by a prior infection with another flavivirus.

The identification of distinct response groups with differing plasmablast expansion rates and maturation dynamics provides important context to our current understanding of the immune response during primary DENV1 infection. Nivarthi et al. (*34*) demonstrated in a controlled human DENV2 infection model with flavivirus-naïve participants that plasmablast expansion, which peaked at day 14, correlated with viral load and that approximately 45% of expanded plasmablast lineages produced DENV2-binding antibodies, the majority of which were serotype-specific. In secondary dengue infections, the surge of DENV-specific and predominantly IgG secreting plasmablasts in the peripheral blood is associated with disease severity (*11*, *13*, *35*).

Taken together, our findings reveal that even under controlled primary DENV1 infection conditions, individuals exhibit distinct B-cell response modalities associated with disease severity. This heterogeneity provides a unique opportunity to dissect the underlying factors driving differential immune activation and may enable the identification of early biomarkers predictive of disease progression, even in primary infections.

### Modelling the B-cell mediated immune response to primary DENV1 infections: cross-reactive memory B-cells, natural antibodies, and impaired affinity maturation

The study participants were pre-screened for antibodies against dengue or Zika NS1 and West Nile Envelope-protein. Serological tests indicate that they raised an IgA, IgG, and IgM response, with the DENV-reactive IgA response occurring around day 16 post-infection (*24*). This observation is reflected in our BCR repertoire analysis, where we detected an early expansion of clones from all three isotypes (Figure 1A), and aligns with earlier findings that the plasmablast isotype distribution is more even in primary than in secondary DENV infections (*10*). The origin of these activated IgA and IgG clones remains unclear. However, their early detection from day 8 (Figure 1B), high SHM rates (Figure 1D), and the lack of temporal correlation in their lineage-tree structures (Supplementary Figure 5) strongly suggests that they are memory-derived. Therefore, the memory-derived clones are either specific to different antigens like other (flavi)viruses, or their serum levels were under the detection threshold prior to infection.

These results indicate that (cross-reactive) memory-derived IgG and IgA plasmablasts form an important compartment of the early B-cell mediated immune response in primary DENV infections, potentially contributing to viral clearance.

Besides the reactivation of memory-derived clones, we observed that primary DENV1 infections lead to the establishment of convergent low *p-gen* IGHM junction sequences during the critical phase of infection (Figure 2A). In the convalescent phase, these same sequences converge in the IGHG fraction while maintaining low SHM rates, strongly suggesting T-cell independent isotype switching as first proposed by Godoy-Lozano et al. (*19*). Indeed, clones expressing these convergent sequences follow the same longitudinal pattern of maturation from IGHM to IGHG (Figure 3).

This aberrant maturation process is a broader property of the immune repertoire, as we found an inverse correlation between the changes in IGHG V-J gene pair expression and SHM rate between day 0 and day 28 (Supplementary Figure 4). The SHM rate reduction is also directly correlated with the ‘publicness’ of clones (Figure 2C) which corroborates prior results by Godoy-Lozano et al. (*19*) and indicates that BCR affinity towards DENV leverages (but is not necessary for) this effect. Our results also agree with those reported by Waickman et al. (*10*) who found that plasmablast IgG sequences exhibited a substantially lower SHM rate than those for IgA in primary DENV infections, although this difference lessens in secondary infections.

Taken together, these results suggest that reduced IGHG SHM rates might be a broader property of the BCR repertoire following DENV infections that, at the level of individual B-cells, is further modulated by BCR specificity.

How could this observation be explained?

It was recently observed that DENV actively replicates in B-cells, with over 40% of B-cells harbouring DENV-RNA during the acute stage (*36*, *37*). These studies likely underestimated the true percentage of infected B-cells, as a DENV infection will affect the health of cells, and dead or dying PBMCs were excluded from analysis.

Ghita et al. (*38*) found that DENV infection causes upregulation of CD69 and CXCR4, which promote antigen internalization and germinal centre homing, respectively. However, the infection also disrupts normal antigen presentation and B-cell receptor signalling by downregulating MHC-II genes (HLA-DPA1, HLA-DRB5) and key BCR-associated genes (FCRLA, SYK, FGR). This impairment of germinal centre interactions may favour T-cell-independent isotype switching over affinity maturation, which aligns with our results.

In parallel, a germinal-centre independent isotype switch could be a conserved antiviral mechanism that allows a fast humoral response. Indeed, Szomolanyi-Tsuda et al. (*39*) found that infectious polyomavirus virions, but not viral antigens, are necessary and sufficient for T-cell independent IgG production in mice. This process can be triggered through viral pattern recognition receptors, such as the Toll-like receptor 3 (TLR3) which recognises dsRNA (*40*), or be regulated through cytokines such as IFN-γ, TNF-α and IL-10 (*37*).

The here presented hypotheses can help explain the overall decrease in SHM of the IGHG repertoires, but they are insufficient to explain why this effect is exacerbated in public, and therefore likely antigen-specific, clones in the convalescent phase.

This aspect could be explained by the recent finding by Gebo et al. (*42*) that DENV-specific BCRs enhance infection of B-cells. Thus, we can hypothesise that antigen-specific clones (which are enriched in public clones) have a higher chance of being infected by the virus and undergo T-cell independent maturation through the aforementioned mechanisms.

The results presented in this study and established literature allow to hypothesise a model for the B-cell mediated immune reaction during primary DENV1 infections (Figure 5).

**Figure 5.**
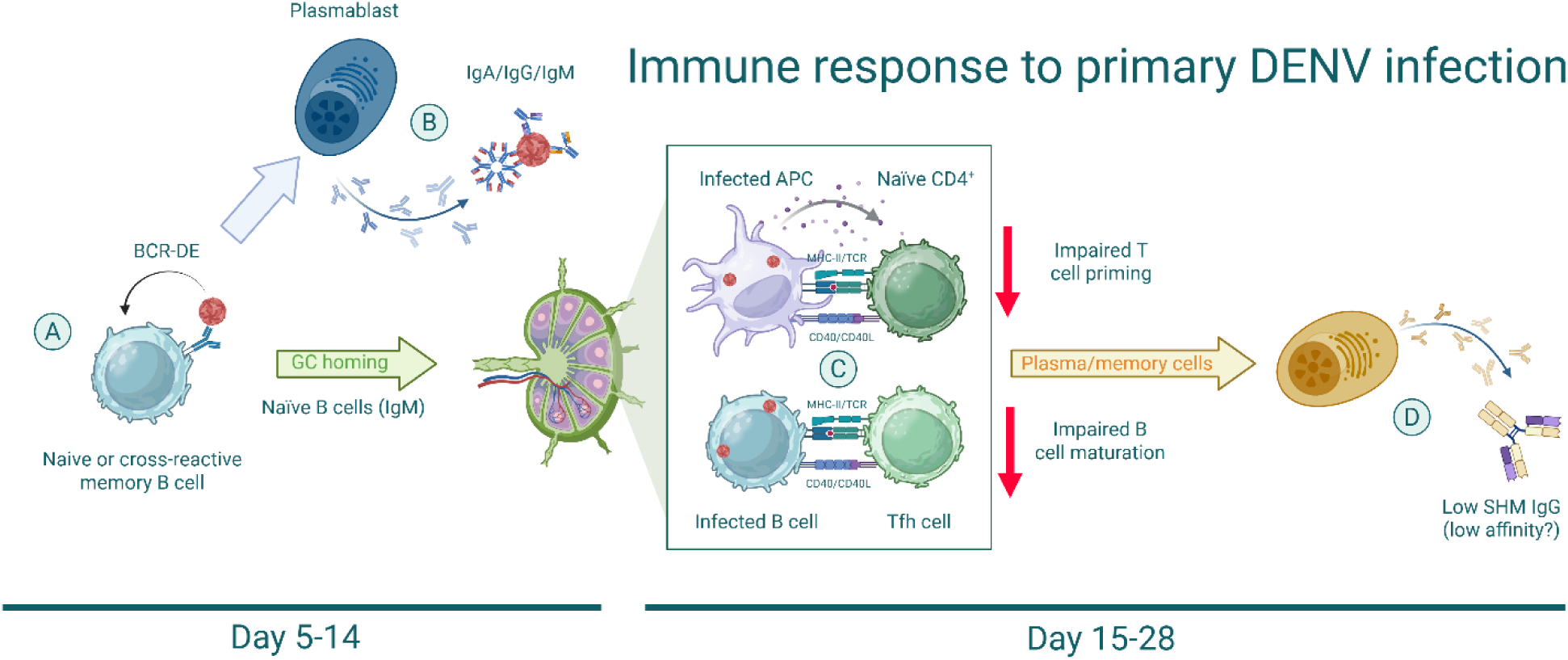
Proposed model for the DENV-mediated dysregulation of the B-cell mediated immune response of primary infections. (**A**) Early in infection, circulating DENV virions interact with the receptors of cross-reactive memory B-cells and reactive naïve B-cells, enhancing their infection. (**B**) During the acute and critical phase, extrafollicular activation leads to rapid expansion of cross-reactive memory B-cells and naïve B-cells into IgA/IgG and germline-like IgM-secreting plasmablasts, respectively. (**C**) DENV infection induces B-cell homing to germinal centres, but impairs affinity maturation via MHC-II downregulation and disrupted APC function. (**D**) In the convalescent phase, germline-like IgG-producing cells emerge, potentially contributing to ADE during secondary infections. Created with BioRender.com.

During the early phase of infection, circulating virions interact with and infect B-cells. This effect is exacerbated in DENV-specific cells, which includes cross-reactive memory B-cells and naïve but reactive B-cells (expressing so-called ‘natural antibodies’), through BCR-dependent enhancement (BCR-DE) (*42*) (Figure 5A).

In the acute phase, cross-reactive memory B-cells are activated and rapidly expand as affinity-matured and predominantly IgA and IgG secreting plasmablasts through the extrafollicular pathway (Figure 1C&D, Supplementary Figure 4, Figure 5B). In the critical phase, naïve B-cells rapidly mature to IgM secreting plasmablasts (Figure 1C) with germline-like receptors leading to the appearance of conserved public junction sequences (Figure 2A). Combined, these processes result in a large release of plasmablasts in the peripheral blood (*13*) secreting affinity matured IgG and IgA antibodies, and germline-like IgM antibodies (Figure 1A, Figure 5B)(*24*).

DENV-infection promotes homing of B-cells to germinal centres for affinity maturation (*37*). DENV impairs the maturation process by downregulating MHC-II expression on B-cells, thereby limiting peptide presentation and cross-signalling with Tfh cells; and by disrupting antigen presentation by antigen-presenting cells (APCs) (Figure 5C)(*37*). This results in the maturation of germline-like IgG producing cells in the convalescent phase, when viral titres in the blood have dropped (Figure 2A&C, Figure 3, Figure 5D)(*24*).

It is intriguing to hypothesize that memory B-cells encoding ‘natural antibodies’, which emerge during a primary infection, may contribute to ADE during secondary infections. This hypothesis is supported by the predominant isotype switching of natural antibodies to IgG (Figure 2A) and the significantly higher IgG fraction observed in secondary infections compared to primary infections (*10*) and subclinical versus hospitalized secondary infections (*17*). Additionally, public junction sequences identified in this manuscript were shown to contribute to the plasmablast pool in hospitalized but not in subclinical secondary DENV infection (*17*).

## Conclusions

This study provides a comprehensive characterization of B-cell repertoire dynamics following primary DENV1 infection in a controlled human challenge model. We identified three distinct response profiles, reflecting fundamental differences in memory B-cell activation, isotype usage, clonal turnover, and maturation dynamics. Across all groups, we observed the emergence of public, convergent B-cell clones during the critical phase that originated from the naïve repertoire. These clones underwent isotype switching to IgG without substantial affinity maturation, pointing toward a T-cell-independent maturation pathway. This delayed, aberrant, or bypassed germinal centre reaction was consistent with prior observations of impaired antigen presentation and MHC-II expression during DENV infection.

### Study limitations and future perspectives

Several constraints may have influenced our findings, and these should be carefully considered when interpreting the data. First, the sampling timepoints were established relative to the moment of infection, whereas most dengue studies typically define day 0 based on symptom onset or cessation. However, it was shown by Waickman et al. (*24*). that viremia peaks occurred at variable intervals among the participants, and the onset of clinical symptoms was likewise not uniform. As a result, direct comparisons with studies using symptom-based timeframes may be limited, and the variability in disease progression can affect our timepoint grouping. For instance, day 10 after viral inoculation corresponded to the start of viremia for certain individuals but the end of viremia for others.

The final sampling point, day 28, may also represent an early phase of convalescence, whereas affinity maturation processes can persist for many months. Consequently, the observed low somatic hypermutation rates of newly identified clones might be confined to this initial stage, potentially overlooking additional clones that develop later. Follow-up studies could incorporate timepoints corresponding more precisely to infection status as well as clinical symptoms to capture ongoing immune maturation.

Second, the exclusive use of whole blood RNA may have skewed sequence data toward plasmablast-derived transcripts, as described by Godoy-Lozano et al. (*19*). Although measures were taken to mitigate potential sampling biases, such as thorough data quality checks and bootstrapping, technical and stochastic limitations cannot be completely ruled out. Greater reliance on cell sorting or single-cell sequencing approaches would likely refine cell-type specificity and improve coverage of less abundant B-cell populations in future work.

Additionally, publicness of novel clones and junctions was used as a surrogate for DENV-specific paratopes. However, this approach cannot definitively confirm DENV specificity, given that some of these clones might also arise towards opportunistic pathogens or other unaccounted for treatment commonalities. Targeted monoclonal antibodies would ideally be cloned and tested *in vitro* to validate dengue reactivity.

Another consideration relates to the under-attenuated DENV strain used in this study, which may not elicit fully representative immune responses compared to those arising from natural infections. Comparative analyses involving both naturally occurring and attenuated lab strains would help determine whether any observed immunological discrepancies are strain dependent. Likewise, only one serotype (DENV1) was included in this study and there might be serotype-specific differences or nuances in the human immune response.

Furthermore, the identification of three convergent clusters induced by DENV infection highlights a need to investigate their functional roles and antigen targets. Studies employing single-cell data and *in vitro* neutralization assays could clarify whether these clusters contribute significantly to protection or, conversely, to processes such as antibody-dependent enhancement. Lastly, while low-SHM clones were identified during primary infections, it remains unknown whether these same clones participate in subsequent responses to different DENV serotypes. Targeted longitudinal sampling and functional testing in secondary infection contexts could establish whether such clones influence disease severity or protection in follow-up exposures.

## MATERIALS AND METHODS

### Study design and sampling

For this study, we made secondary use of biobanked total RNA samples originally collected in the context of a controlled human dengue virus challenge trial (*24*). The RNA samples were extracted from whole blood, as previously described in the original clinical study. For the present analysis, we included samples from nine participants and five timepoints: days 0, 8, 10, 14, and 28 post-infection. The timepoints were classified based on the DENV1 RNA content in the serum reported by Waickman et al. (*24*): the viral titers were first detectable at (mean ± SD) day 7.8 ± 1.5, peaked at day 10.6 ± 1.5, and were last detectable at day 14 ± 1.8. Based on these results, the timepoints are classified as shown in Table 1.

**Table 1.** Naming of sampling timepoints based on published virology and serology data.

| Timepoint (days) | Classification |
| --- | --- |
| 0 | Pre-infection |
| 8 | Early viremic |
| 10 | Acute / peak viremic |
| 14 | Critical / late viremic |
| 28 | Convalescent |

### RepSeq library preparation, quantification, and sequencing

The RNA concentration was measured using the Qubit^TM^ High Sensitivity RNA Assay (Thermo Fisher Scientific). All samples were further concentrated to ≥200 ng/μL using the Zymo RNA Clean & Concentrator™-5 kit (Zymo Research) and quantified using the Qubit^TM^ High Sensitivity RNA Assay. Samples day 0 of participant 6; and day 28 of participant 1; were not processed due to too low input RNA availability. For library preparation, the NEBNext® Immune Sequencing Kit (New England Biolabs) was employed. First, cDNA was produced from 1 μg of total RNA following manufacturer’s instructions. Next, the cDNA was split into three equal parts and used as template for the PCR1 reaction using the NEBNext® Immune Sequencing Primers (either BCR or TCR, product code E2629L, New England Biolabs) or IGH-specific primers (Supplementary Table 2) using 2/3^rd^ reaction volumes for each prep. The optional qPCR quantification step was used to ensure sufficient amplification of the IGH libraries, which required up to five time more cycles than the BCR reaction of the same sample. The final library sizes were verified using a High Sensitivity D1000 ScreenTape on the 4150 TapeStation system (Agilent) following manufacturer’s instructions.

The libraries were quantified through Quantitative Real-Time PCR on a CFX96^TM^ system (Bio-Rad). Reaction mixes were prepared by combining 1 µL of 1/2000 diluted libraries (in 0.1% Tween-20) with 5 µL of KAPA SYBR FAST Master Mix Universal (Merck), 0.1 µL of forward (5’-AATGATACGGCGACCACCGAGAT-3’) and reverse (5’-CAAGCAGAAGACGGCATACGA-3’) primers at 10 µM each, and 3.8 µl water. Each sample was run in triplicate. Libraries were quantified relative to NEBNext® Library Quant DNA Standards (New England Biolabs).

Libraries were pooled equimolarly to a concentration of 10 µM and 24 µl was used as input for the Element Adept Library Compatibility Kit v1.1 (Element Biosciences) following manufacturer’s instructions. The circularized libraries were sequenced on an AVITI system (Element Biosciences) using 300PE kit and medium output flow cell with 30% PhiX.

### Bioinformatics

Analyses were carried out in R v4.4.1 (*43*) supported by packages from the tidyverse (*44*), Immcantation suite, and python v2.7.

### Sequence assembly and clone determination

Raw sequencing reads in FASTQ format were processed using the nf-core/airrflow analysis pipeline (*45*) version 4.1.0 using the *nebnext_umi* protocol. The total count of unique BCR heavy-chain sequences is shown in Supplementary Table 1. For clonal analysis of the BCR libraries, the clustering threshold Hamming distance value was first estimated automatically through the pipeline and then verified manually. When the computed threshold value did not separate the two modes of the distance-to-nearest distribution, it was set manually and the clonal analysis rerun.

### Public clone mutation frequency

The IGH sequences from all participants were first pooled and clustered using single-linkage hierarchical clustering. This clustering was based on V-J gene pairing and a length-normalized CDR3 amino-acid Hamming distance threshold of 0.2, using the *hierarchicalClones()* function from the SCOPer R package (*46*). The number of unique subjects contributing to each cluster was defined as the ‘cluster subject count’. Clones containing sequences belonging to a cluster with subject count over one were marked as public clones. To analyse the correlation between clone publicness and mutation frequencies over time across different IGH isotypes (IGHA, IGHG, and IGHM), sequences were filtered to retain only these isotypes. The repertoires were then aggregated at the level of subject, clone, collection time point, and isotype; computing the mean mutation frequency within each grouping and defining the publicness of the grouping as the highest ‘cluster subject count’ of any of its constituent sequences. Next, for each isotype and time point, the mean mutation frequency and its standard deviation for each level of publicness were computed.

### Clone tracking analysis

Assembled BCR repertoires were filtered for IGHA, IGHG, and IGHM sequences. The sequences were grouped by subject and timepoint, then the complete clone RAD was computed through the *estimateAbundance()* function as previously described (*47*). For each grouping, the 200 most abundant clones were determined and their abundance tracked. Repertoire overlap metrics (Jaccard (*48*) and Morisita-Horn (*49*) indices) were computed between days 0-8, 8-14, and 14-28; and the mean value calculated.

### Public junction analysis and sequence logos

The generation probabilities of all junction amino acid sequences shared by two or more participants were inferred with OLGA version 1.2.4 using the default human B-cell heavy chain model (VDJ) (*50*). To identify public junction clusters, an adjacency matrix was generated with all identified public junctions through the *stringdist* R package version 0.9.12 (*51*) setting a Levenshtein distance threshold of 2. This metric and threshold were chosen as they allow to cluster functionally similar CDR3s (*52*). Distinct clusters were then identified through the *igraph* R package version 2.1.1 (*53*). To generate sequence logos of identified clusters, for each cluster the junction amino acid sequences were aligned with the *msa* R package version 1.38.0 (*54*) using the ClustalW method. Alignments were then visualized with the *ggseqlogo* R package version 0.2 (*55*).

### Phylogenetic trees and time correlation analysis

The dowser trees were generated by the nf-core/airrflow pipeline and processed with custom scripts to add time point, duplicate count, and junction amino acid sequence traits to each node. Trees with nodes from less than two time points or with any node from collection time point 0 were filtered out. A time correlation test was performed with the *correlationTest()* function from the dowser R package version 2.3 (*56*). The phylogenetic trees were rendered with the *ggtree* R package version 3.14.0 (*57*).

### V-J gene family usage analysis

Differences in V-J gene family pair usage over time (days 0, 14, and 28) were evaluated with a zero-inflated negative binomial mixed-effects model implemented with the *glmmTMB* package version 1.1.10 (*58*) in R. The repertoires were filtered to include only the IGHA, IGHG, and IGHM sequences, then the model was applied to raw counts of V-J gene pair occurrences, grouped by collection time point, isotype, public/private clone status, and subject. The response variable was the raw count of each gene pair, with collection time point as a fixed effect and subject as a random effect to account for inter-individual variability. An offset term, defined as the logarithm of the total sequence count per group, was included to normalize for differences in sequencing depth. Estimated marginal means and contrasts (day 0 vs. 14 and day 0 vs. 28) were computed using the *emmeans* package version 1.10.7 (*59*) to derive p-values. The p-values were adjusted for multiple comparisons through a Benjamini–Hochberg correction and significance defined as adjusted p < 0.05 and absolute log2 fold change > 1.

### Graphics

Plots were generated through custom scripts using the ggplot2 R package (*60*).

## Supplementary Materials

- Supplementary Figure 1 – Clone ranking and diversity plots.
- Supplementary Figure 2 – Temporal effects of IGH size.
- Supplementary Figure 3 – Gene-pair usage volcano plots.
- Supplementary Figure 4 – Correlation between V-J gene pair usage and SHM rate.
- Supplementary Figure 5 – Phylogenetic trees of largest clones.
- Supplementary Table 1 – Count of unique IGH sequences identified.
- Supplementary Table 2 – Human primers E2628L.

## Data Availability

All data produced in the present study will be available upon publication

## Funding

Agency of Innovation and Entrepreneurship of the Flemish Government HBC.2022.0988 Johnson and Johnson

State of New York

Congressionally Directed Medical Research Program

## Author contributions

Conceptualization: TWV, ASV, CS, FVN, OL, KKA

Methodology: TWV, MS

Investigation: TWV, MS

Visualization: TWV

Funding acquisition: FVN, AW, OL, KKA

Project administration: FVN, AW, OL, KKA

Supervision: FVN, OL, KKA

Writing – original draft: TWV

Writing – review & editing: TWV, ASV, CS, MS, FVN, AW, OL, KKA

## Data and materials availability

Upon publication in a peer-reviewed journal, the computational pipeline employed in this paper will be published as a GitHub repository under: https://github.com/TWV-GIT/science_immunology_J9_AIRR.

The raw sequencing reads will be archived on the NCBI SRA database.

## Footnotes

^1^A clone is defined as a group of cells that share a common naïve B-cell progenitor. This definition is synonymous with lineage.

