## Supplementary Materials for "Establishment of ‘natural antibodies’ during primary dengue infection"

##### Supplementary Figure 1: clone ranking and diversity plots

Important metrics of BCR and TCR immune repertoires are the total count of unique clones sampled (termed 'species richness') and the proportional contribution of these clones to the entire repertoire. Combined, these parameters allow to calculate Hill diversity profiles (*27*) that can accurately predict immune status (*28*). Clone ranking and repertoire evenness were assessed for each participant through the alakazam package version 1.3.0 (*49*). First, the complete species-rank abundance distribution (RAD) of clones at each timepoint was imputed for the IGHG, IGHA, and IGHM fractions separately through the one-parameter estimator function *estimateAbundance()* (*26*). Because the diversity values are sensitive to technical and biological sampling (*50*), the RADs were inferred through resampling with 200 bootstrap realizations as to derive confidence intervals (*51*) using the alakazam library (*49*). Abundances were estimated considering the copy number of unique sequences. Repertoire diversity curves were then computed using the generalized diversity index (*27*) function *alphaDiversity()*. The repertoire evenness was derived by dividing the Renyi entropy by the species richness ${}^{q}E(f) = \frac{(\sum_{i=1}^{n} f_i^{q})^{\frac{1}{1-q}}}{n}$.

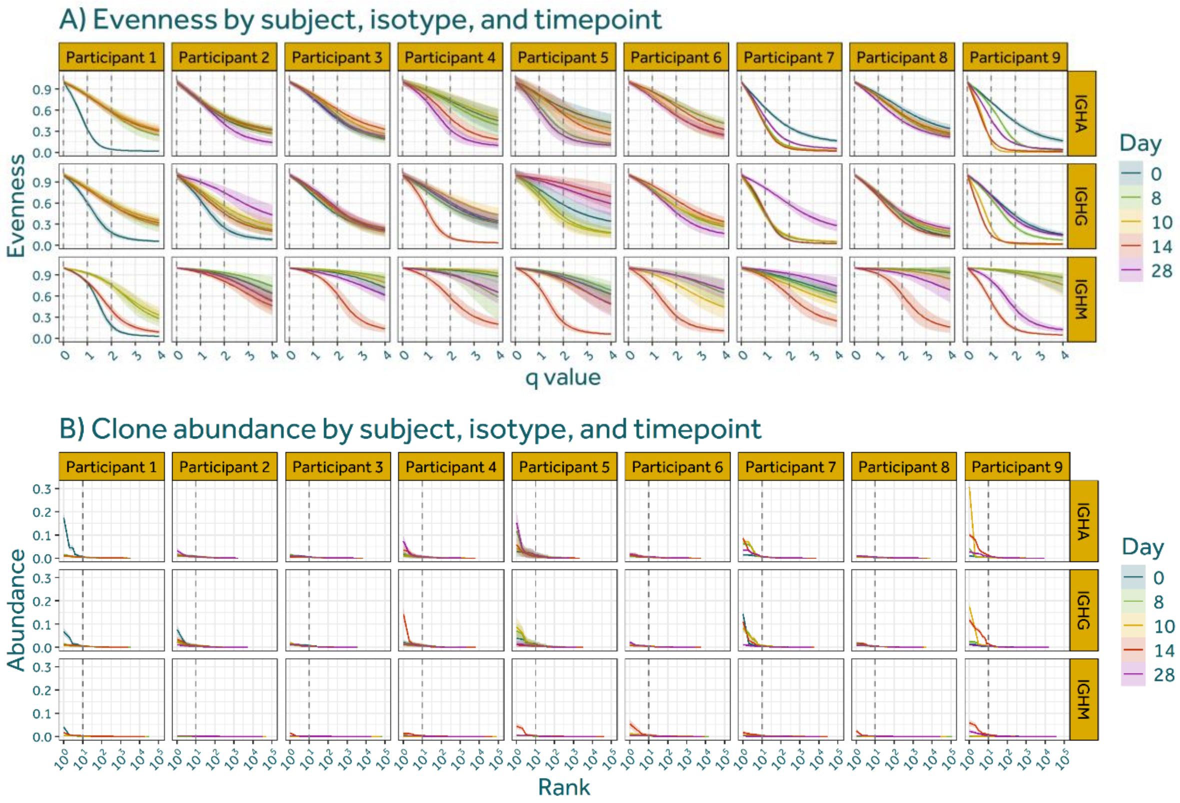


*Supplementary Figure 1 –* ***Clone ranking and diversity plots.*** *(**A**) Repertoire evenness curves for IGH sequences split by isotype and timepoint. Shaded areas indicate 95% confidence intervals. (**B**) Ranking of clones by relative abundance, split by isotype and day.*

The evenness curves (Supplementary Figure 1A) show that the primary DENV1 infection induced a strong activation of the IGHM compartment on day 14 after inoculation for all participants except 1 and 2; In participant 1 this trend might have been broken due to an exceptionally low repertoire diversity at timepoint 0. The response of the IGHG and IGHA isotypes are highly participant-dependent, both in strength and timepoint.

The clone-rank plot (Supplementary Figure 1B) shows marked differences in the degree of clonal expansion between participants: participants 1 and 7 each show a hyperexpanded clone at timepoint 0 (18% of IGHA sequences and 14% of IGHG sequences respectively), indicative of an active B-cell response prior to DENV1 experimental challenge.

**Supplementary Figure 2: temporal effects of IGH size**

The Shannon entropy of IGH, IGK, and IGL CDR3 lengths was calculated as previously described (*19*). In summary, sequences were filtered with a CDR3 sequence length from 4 to 34 amino acids (inclusive). As the *hierarchicalClones()* function from the SCOPer R package version 1.3.0 does not allow to infer clonal lineages from light-chain sequences of bulk data (*52*), new clonotypes were defined for each locus as unique V-J gene combinations with identical CDR3 amino acid sequence (*19*). To ensure comparable diversity estimates across samples with different sequencing depths, datasets were subsampled to an equal unique sequence count for each sample and locus (n = 1846). This standardization controls for the fact that deeper sequencing can detect additional rare clonotypes that would be missed in smaller samples, which would otherwise confound comparisons of Shannon entropy between samples.

To evaluate temporal changes in the distribution of IGH CDR3 lengths, weighted mean CDR3 lengths were computed per subject using the count of each length as a weight. Statistical comparisons across time points were performed separately for each isotype using linear mixed-effects models with time point as a fixed effect and subject as a random intercept. The reference level was set to day 0. Post-hoc contrasts comparing each subsequent time point to day 0 were conducted using estimated marginal means with Sidak correction for multiple testing. Adjusted p-values $< 0.05$ were considered significant.

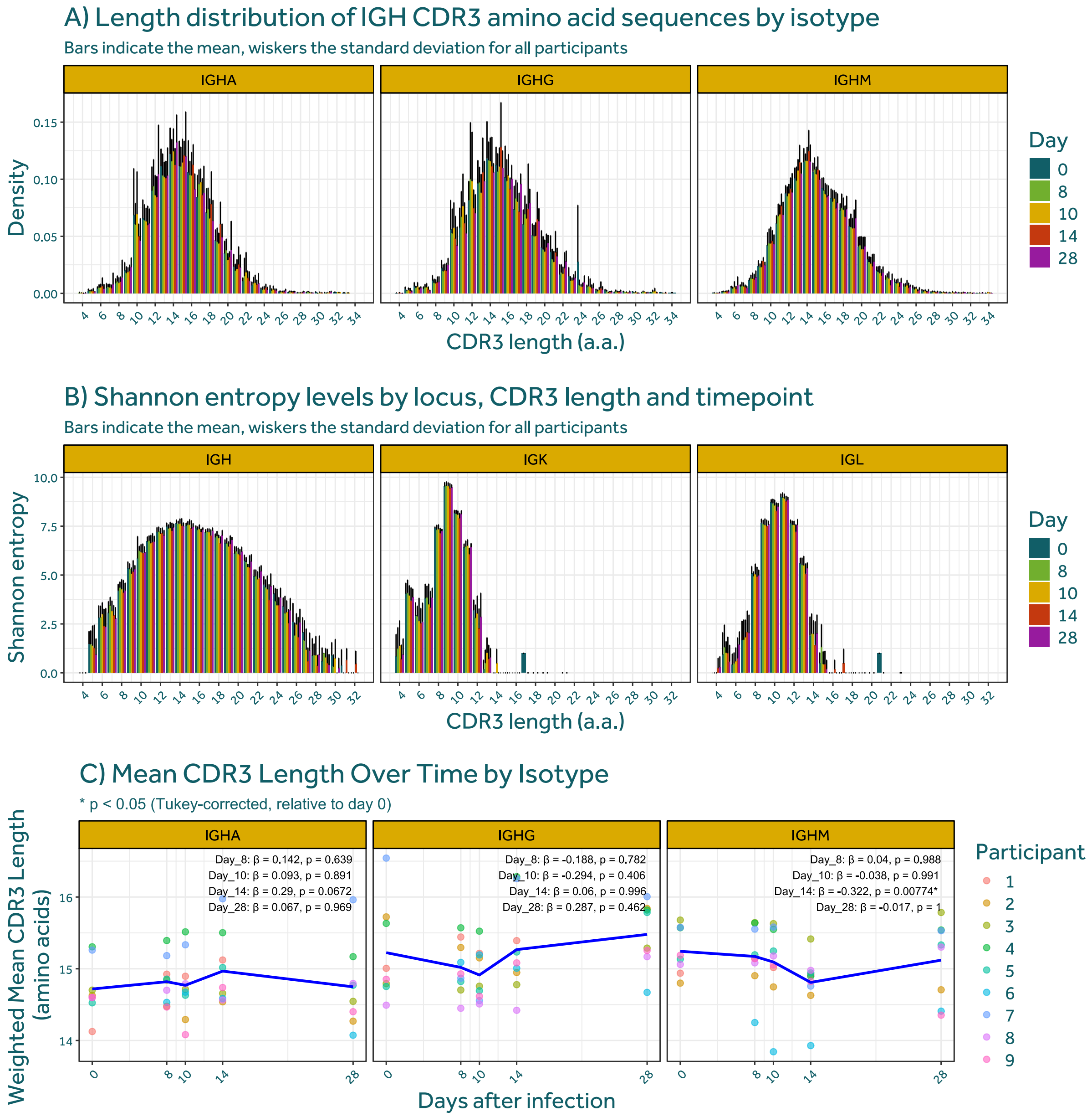


*Supplementary Figure 2 -* ***Temporal effects of IGH size****. (****A****) Density distribution of IGH CDR3 lengths split by isotype and timepoint. (****B****) Shannon entropy of IGH CDR3 lengths split by isotype and timepoint. Bars indicate the mean of all participants, whiskers the standard deviation. (****C****) Weighted mean CDR3 lengths (in amino acids) were calculated per participant, time point, and isotype (IGHA, IGHG, IGHM). Individual points represent participants; blue lines indicate group means over time. Statistical comparisons were performed using linear mixed-effects models with time as fixed effect and subject as random effect. Post-hoc contrasts vs. day 0 were corrected using the Sidak method. Asterisks (*) denote statistically significant differences (adjusted p < 0.05).*

#### Supplementary Figure 3: gene-pair usage volcano plots.

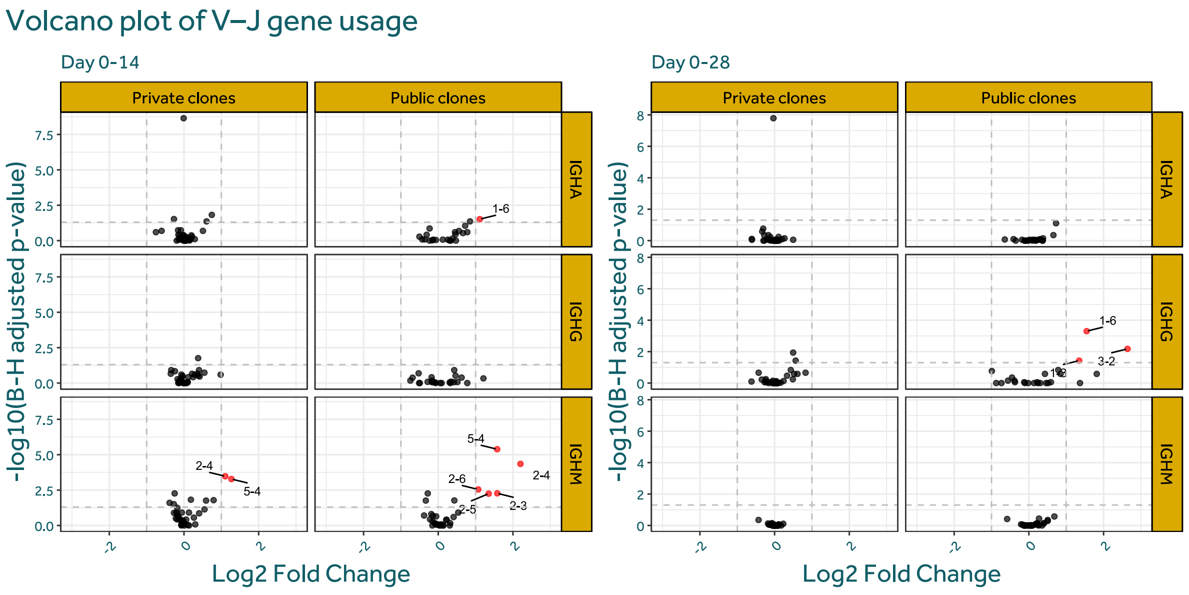


*Supplementary Figure 3 -* ***Gene-pair usage volcano plots****. Volcano plots for the change in gene-family pair usage between timepoints 0-14 and 0-28. Significantly changed gene-family pairs coloured in red. P-values calculated through a zero-inflated negative binomial mixed model with Benjamini–Hochberg correction.*

**Supplementary Figure 4 – Correlation between V-J gene pair usage and SHM rate.** The relationship between changes in somatic hypermutation (SHM) frequency and V–J gene-pair usage over time were analysed. The differential log2 fold change gene-expression and absolute SHM change (day 14 and 28 relative to baseline day 0) data plotted in Figure 4 were used. Pearson correlation coefficients were calculated for each isotype, public clone status, and time point combination. Correlation p-values were obtained using cor.test() and adjusted for multiple testing using the Benjamini-Hochberg method.

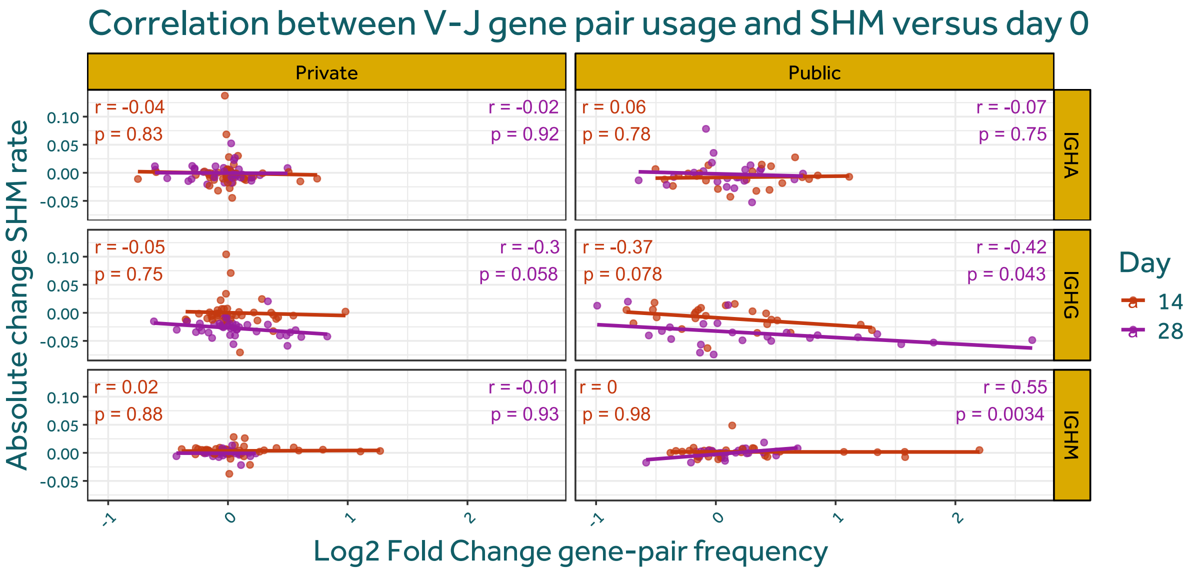


*Supplementary Figure 4 –* ***Correlation between V-J gene pair usage and SHM rate****. Each point represents a gene-family pair, with colours denoting the timepoint (day 14 or 28). SHM differences (y-axis) are compared against log2 fold changes in gene-pair frequency (x-axis). Linear regression lines are overlaid per timepoint. Pearson correlation coefficients (r) and associated p-values are annotated for each facet and timepoint. p-values were adjusted using the Benjamini-Hochberg method to control for multiple testing.*

### Supplementary Figure 5 – Phylogenetic trees of largest clones

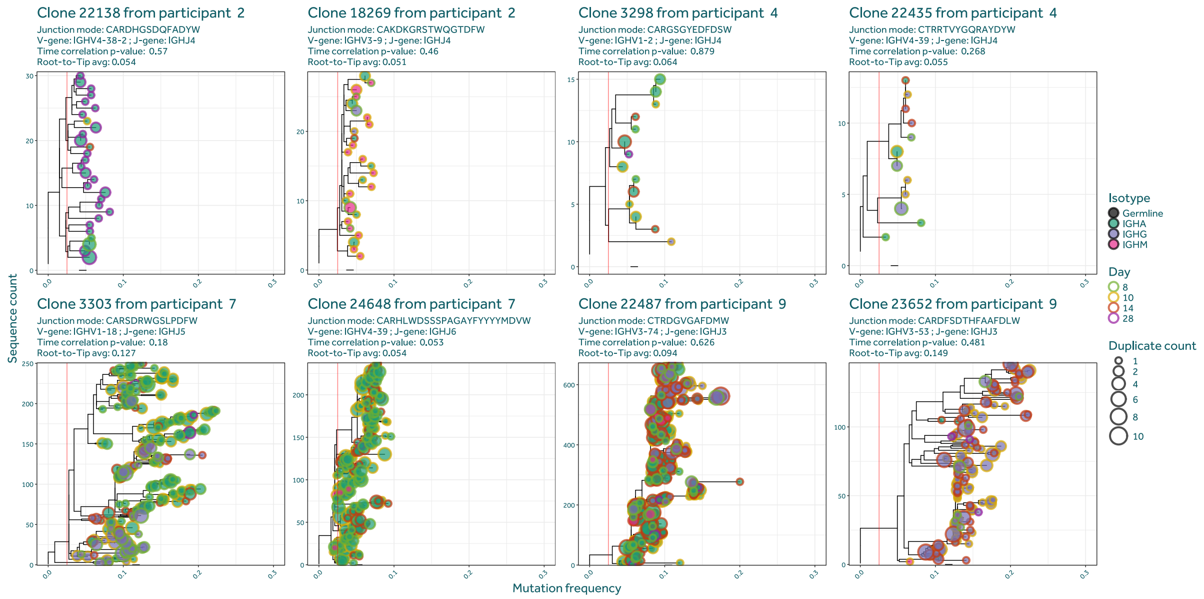


*Supplementary Figure 5 – **Phylogenetic trees of largest clones**. Trees for the two largest clones from each participant that were absent at timepoint 0 and are observed at timepoints 8 and 10. Trees were generated by the nf-core/airrflow and further processed with custom scripts. Size of the nodes is cut off at count = 10.*

#### Supplementary Tables

*Supplementary table 1* - ***Count of unique IGH sequences identified.*** *Row headers indicate the participants, column headers the collection timepoint.*

| ID | **Collection day** | | | | |
|---|---|---|---|---|---|
| | **0** | **8** | **10** | **14** | **28** |
| **1** | 7452 | 7933 | 5533 | 8335 | NA |
| **2** | 8655 | 9107 | 8313 | 3748 | 7146 |
| **3** | 8193 | 13432 | 10571 | 9131 | 5639 |
| **4** | 5712 | 3913 | 5948 | 15073 | 2766 |
| **5** | 2352 | 4026 | 1846 | 24083 | 2350 |
| **6** | NA | 7425 | 3835 | 8961 | 3271 |
| **7** | 5392 | 5227 | 3960 | 25277 | 7713 |
| **8** | 10073 | 11868 | 11869 | 5906 | 5535 |
| **9** | 10294 | 12919 | 12743 | 14712 | 18345 |

*Supplementary table 2* - ***Human primers E2628L***

| | | | |
|---|---|---|---|
| BCR (NEBNext) | IGH | IGH$_M$ | ACACTCTTTCCCTACACGACGCTCTTCCGATCTGAATTCTCACAGGAGACGAGG |
| | | IGH$_D$ | ACACTCTTTCCCTACACGACGCTCTTCCGATCTTGTCTGCACCCTGATATGATGG |
| | | IGH$_A$ | ACACTCTTTCCCTACACGACGCTCTTCCGATCTGGGTGCTGYMGAGGCTCAG |
| | | IGH$_E$ | ACACTCTTTCCCTACACGACGCTCTTCCGATCTTTGCAGCAGCGGGTCAAGG |
| | | IGH$_G$ | ACACTCTTTCCCTACACGACGCTCTTCCGATCTCCAGGGGGAAGACSGATG |
| | IGL | IGL$_K$ | ACACTCTTTCCCTACACGACGCTCTTCCGATCTGACAGATGGTGCAGCCACAG |
| | | IGL$_L$ | ACACTCTTTCCCTACACGACGCTCTTCCGATCTAGGGYGGGAACAGAGTGAC |
| TCR (NEBNext) | | TR$_A$ | ACACTCTTTCCCTACACGACGCTCTTCCGATCTCACGGCAGGGTCAGGGTTC |
| | | TR$_B$ | ACACTCTTTCCCTACACGACGCTCTTCCGATCTCGACCTCGGGTGGGAACAC |
| | | TR$_D$ | ACACTCTTTCCCTACACGACGCTCTTCCGATCTCGGATGGTTTGGTATGAGG |
| | | TR$_G$ | ACACTCTTTCCCTACACGACGCTCTTCCGATCTGGGAAACATCTGCATCAAG |
